# Epidemiological, clinical and genomic insights into the ongoing diphtheria outbreak in Yemen

**DOI:** 10.1101/2020.07.21.20159186

**Authors:** Edgar Badell, Abdulilah Alharazi, Alexis Criscuolo, The NCPHL diphtheria outbreak working group, Noemie Lefrancq, Valerie Bouchez, Julien Guglielmini, Melanie Hennart, Annick Carmi-Leroy, Nora Zidane, Marine Pascal-Perrigault, Manon Lebreton, Helena Martini, Henrik Salje, Julie Toubiana, Fekri Dureab, Ghulam Dhabaan, Sylvain Brisse

**Affiliations:** Institut Pasteur, Biodiversity and Epidemiology of Bacterial Pathogens, Paris, France; Institut Pasteur, National Reference Center for Corynebacteria of the diphtheriae complex, Paris, France; National Centre of the Public Health Laboratories (NCPHL), Sana’a, Yemen; Hub de Bioinformatique et Biostatistique ⍰ Département Biologie Computationnelle, Institut Pasteur, USR 3756 CNRS, Paris, France; Mathematical Modelling of Infectious Diseases Unit, Institut Pasteur, UMR2000, CNRS, Paris, France; Sorbonne Université, Collège doctoral, F-75005 Paris, France; Department of Microbiology, National Reference Centre for toxigenic corynebacteria, Universitair Ziekenhuis Brussel, Vrije Universiteit Brussel (VUB), Laarbeeklaan 101, 1090 Brussels, Belgium; Department of Genetics, University of Cambridge, Cambridge, UK; Université de Paris, General Paediatrics and Paediatric Infectious Diseases Department, Hôpital Necker-Enfants malades, APHP, Paris, France; Institute for Research in International Assistance, Akkon Hochschule, Berlin, Germany, 2-Heidelberg Institute of Global Health, Heidelberg, Germany; Mount Sinai Hospital, University Health Network, University of Toronto, Toronto, Canada

**Keywords:** diphtheria, outbreak, epidemiology, phylogenetics, antimicrobial resistance, genomic sequencing, evolutionary rate, Yemen

## Abstract

**Background:** An outbreak of diphtheria, declared in Yemen in October 2017, is still ongoing. Methods. Probable cases were recorded through an electronic diseases early warning system. Microbiological culture, genomic sequencing, antimicrobial susceptibility and toxin production testing were performed.

**Methods:** Probable cases were recorded through an electronic diseases early warning system. Microbiological culture, genomic sequencing, antimicrobial susceptibility and toxin production testing were performed.

**Findings:** The Yemen diphtheria outbreak developed in three epidemic waves, which affected nearly all governorates (provinces) of Yemen, with 5701 probable cases and 330 deaths (October 2017 - April 2020). The median age of patients was 12 years (range, 0.17-80). Virtually all outbreak isolates (40 of 43 tested ones) produced the diphtheria toxin. We observed low level of antimicrobial resistance to penicillin. We identified six separate *Corynebacterium diphtheriae* phylogenetic sublineages, three of which are genetically related to isolates from Saudi Arabia and Somalia. The predominant sublineage was resistant to trimethoprim and was associated with unique genomic features, more frequent neck swelling (*p*=0.002) and a younger age of patients (*p*=0.06). Its evolutionary rate was estimated at 1.67 × 10^−6^ substitutions per site year^-1^, placing its most recent common ancestor in 2015, and indicating silent circulation of *C. diphtheriae* in Yemen earlier than outbreak declaration.

**Interpretation:** We disclose clinical, epidemiological and microbiological characteristics of one of the largest contemporary diphtheria outbreaks and demonstrate clinically relevant heterogeneity of *C. diphtheriae* isolates, underlining the need for laboratory capacity and real-time microbiological analyses to inform prevention, treatment and control of diphtheria.

**Funding:** This work was supported by institutional funding from the National Centre of the Public Health Laboratories (Sanaa, Yemen) and Institut Pasteur (Paris, France) and by the French Government Investissement d ‘Avenir Program.

## Introduction

Diphtheria is a severe infection that typically affects the upper respiratory tract, potentially leading to pseudomembrane formation, neck swelling and suffocation [1, 2]. In addition, diphtheria toxin production by some *C. diphtheriae* strains can cause damage to the heart and other organs. Non-toxigenic strains can also cause invasive infections [3]. Before large scale vaccination, which mainly targets the toxin and is highly effective in preventing the disease, diphtheria was a major cause of death in children [2]. Large outbreaks of diphtheria are often observed following disruption to vaccination programs, as occurred in the Soviet Union in the 1990s [4] and more recently in the Rohingya refugee population in Bangladesh [5] and in Venezuela [6]. Diphtheria is still observed occasionally in high-income countries, in particular among migrants [7, 8].

In Yemen, where civil war has been raging since March 2015, a large outbreak of diphtheria was recognized in October 2017 [9, 10][11]. While a vaccination campaign targeting 300,000 children began in late November 2018, with plans to scale up to 3 million children and young adults in December, the ongoing conflict has complicated vaccination catch-up efforts [12, 13]. Before this outbreak, diphtheria was considered endemic in Yemen, with an average of 50 suspected cases reported annually since 2000 [14]. The last documented outbreak of diphtheria in this country occurred in 1981-82, with a total of 149 cases with no death. Yemen is currently also affected by other epidemic diseases including cholera [15] and COVID-19 [16, 17]. As of May 2020, the diphtheria outbreak was still ongoing [18]. The early epidemiology of the outbreak was described [9, 10], but virtually no data have been reported on the clinical and microbiological characteristics of the outbreak.

Besides vaccination, a critical component of diphtheria management is serotherapy. Unfortunately, diphtheria antitoxin is not readily available in Yemen, as in most world countries [19]. Antimicrobial treatment is also indicated for patient care and to avoid transmission, with penicillin and erythromycin being recommended as first-line therapeutic agents [20][2].

Here we report on the clinical, epidemiological and microbiological features of the ongoing Yemen diphtheria outbreak.

## Results

### Epidemiology and clinical features of the Yemen diphtheria outbreak, 2017-2020

Large numbers of probable cases were recorded in 2017 (n=560), 2018 (n=2606), 2019 (n=2004) and in 2020 (up to week 17: n=649). Altogether, 5701 cases including 330 deaths were reported up to April 26th, 2020, corresponding to a case fatality rate of 5.8%. Between 2017 and 2020 there were three major waves of cases (**Figure 1A**). The first wave occurred from October 2017 until June 2018 with a peak on week 6 of 2018. The second wave was flatter and lasted from June 2018 until June 2019. The third wave started in June 2019 and reached its peak in October 2019, with more than 80 cases/week by that time.

**Figure 1.**
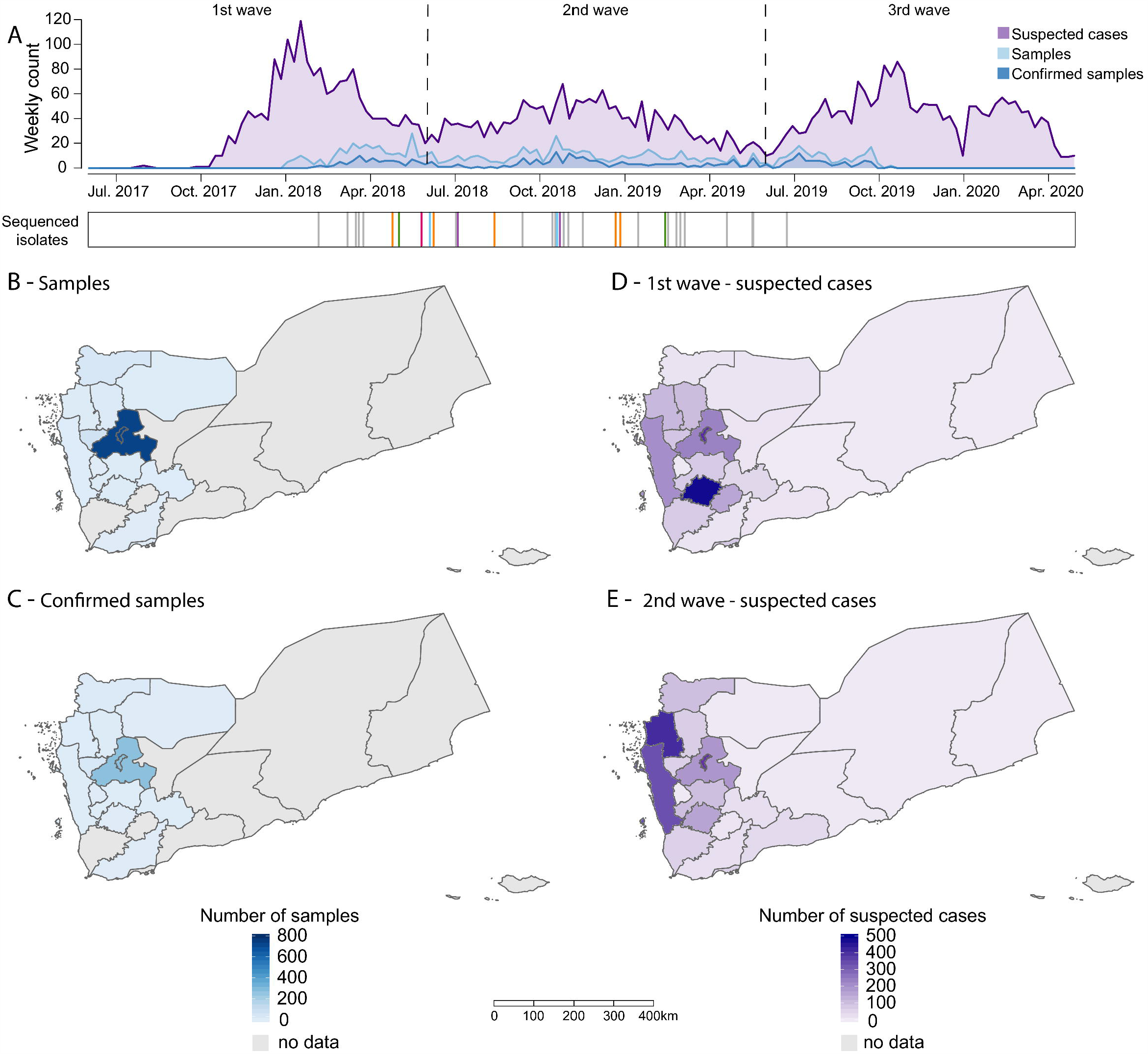
Epidemic curve and geographic and temporal origins of the isolates. A. Weekly numbers of suspected cases (purple), samples (light blue) and confirmed isolates (dark blue) of diphtheria in Yemen from January 2017 to April 2020, with three epidemic waves being highlighted. Sequenced isolates are shown below the epidemic curve, colored according to their sublineages (see Figure 3). B and C. Geographical location of the 888 samples of this study (B) and of the 340 confirmed samples (C). D and E. Geographical location of the suspected cases during the first epidemic wave (D) and the second epidemic wave (E). The governorate maps of Yemen was created using a shape file approved by the Humanitarian Country Team in October 2019 (https://data.humdata.org/dataset/yemen-admin-boundaries).

The outbreak affected nearly all governorates of Yemen (**Figure 1, Table S3**). However, some governorates were more affected and the two first epidemic waves had distinctive geographic signatures (**Figure 1D,E**). The outbreak started in October 2017 in Ibb, where 288 cases (52% of the total 2017 cases) were registered; while Al Hodeidah recorded 60 cases (11%) in this year. In 2018, 2606 cases were registered in 21 governorates, with 64% of the cases registered in 5 governorates: Hajja (395 cases, 15%), Al Hodeidah (338, 13%), Sana ‘a (339, 13%), Ibb (324, 12%) and the capital city of Sana ‘a (276, 11%). Last, the distribution of reported cases in 2019 shows that 60% of cases occurred in four governorates: Al Hodeida (371 cases, 18%), Hajjah (327 cases, 16%), Sadah (297 cases, 15%) and Sana ‘a (226 cases, 11%).

Clinical data available for 888 patients (February 2018 - November 2019) showed that patients were more likely to be female (55%), and their median age was 12 (range, 0-80) years (**Figure 2A**). Of the 750 patients with available data, 434 (58%) reported a history of previous vaccination against diphtheria. The vaccinated proportion of patients did not change between 2018 and 2019 (57% vs. 59%, p=0.49). The median age of vaccinated patients was lower (10 years, range 0.17-45) than that of non-vaccinated patients (13 years, range 0.17-80; p<0.001). The age distribution of cases was available for the first two epidemic waves. We found that the median age of infection was 12 years old; however, the attack rate was highest in those 5-15 years in age as compared to other groups. While the attack rate was largely constant between the first two years in those over 5 years old, there was a marked reduction in children 0-4 years between the two waves (**Figure 2B; Figure S1**). Almost all patients (n=818/841, 97%) had fever. Pseudomembrane was present in 787/839 (94%), signs of laryngitis in 548/784 (31%), neck swelling in 456/769 (59%) and tonsillitis in 821/836 (98%) patients with available data. Difficulty to swallow and to breathe were observed in 710 (86%) and 399 (51%) patients, respectively. No difference in disease expression was observed between vaccinated and non-vaccinated patients. Most patients (n=634/790, 80%) were reported to be under antibiotic therapy at the time of throat sampling.

**Figure 2.**
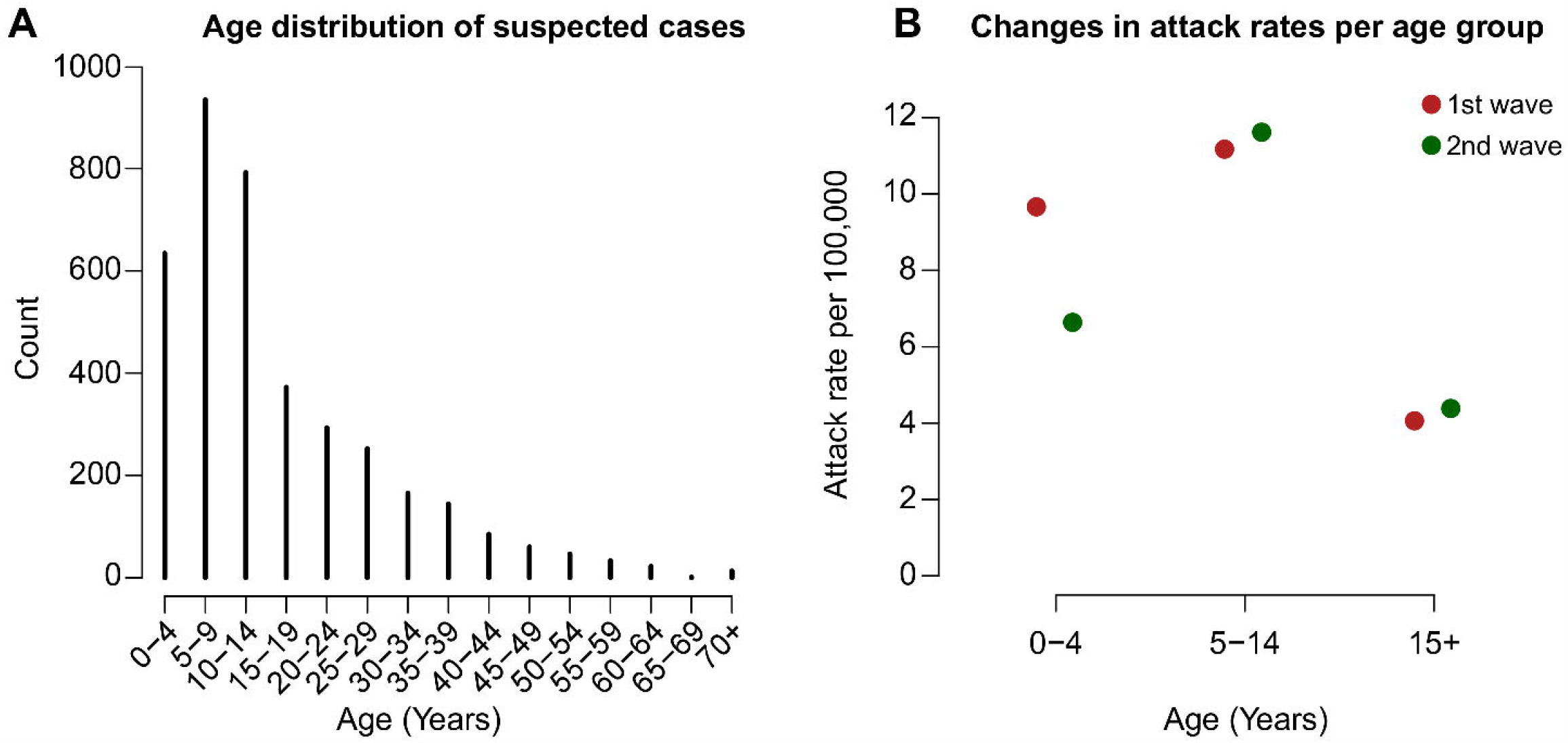
Age profile of suspected cases. A. Age distribution of suspected cases during the first and second waves. B. Attack rates per age group, per wave. Red shows the first wave, green the second. The mean attack rate during the two waves was 6.6 per 100,000.

Microbiological confirmation of cases was tested for 888 samples referred to the NCPHL from 2018 to 2020. Of the 836 cases with available results, 340 (41%) were microbiologically confirmed by a positive *C. diphtheriae* culture. The temporal and geographic distributions of tested and confirmed cases were similar (**Figure 1B-C**).

### Phylogenetic diversity of *C. diphtheriae* from Yemen and their relationships with global isolates

A random selection of 98 *C. diphtheriae* positive cultures were transferred to Institut Pasteur (Paris, France) for external confirmation and sequencing. Two cultures (YEM0065 = YE-NCPHL-647L) and YEM0070 = YE-NCPHL-647S) with distinct colony morphologies had been isolated from a single sample. We confirmed by qPCR [21] that 47 cultures carried genetic material of *C. diphtheriae*, 43 of which could be isolated in pure culture.

Detection of the diphtheria toxin *tox* gene was positive in 40 out of 43 isolates; 3 isolates were *tox*-negative by qPCR (**Table S1**), which was confirmed by genomic analyses. The Elek *in-vitro* test for diphtheria toxin production demonstrated that 39 *tox*-positive isolates produced the toxin (YEM0035 was Elek negative).

Phylogenomic analyses revealed the existence of six sublineages (labeled A to F, **Figure 3**). One of them, called sublineage A, comprised 30 (70%) isolates (**Table S1**). The six sublineages were unrelated to each other, as evidenced by their scattered positions in a global phylogenetic tree of *C. diphtheriae* (**Figure S2**), demonstrating multiple occurrences of diphtheria re-emergence or introductions into Yemen.

**Figure 3.**
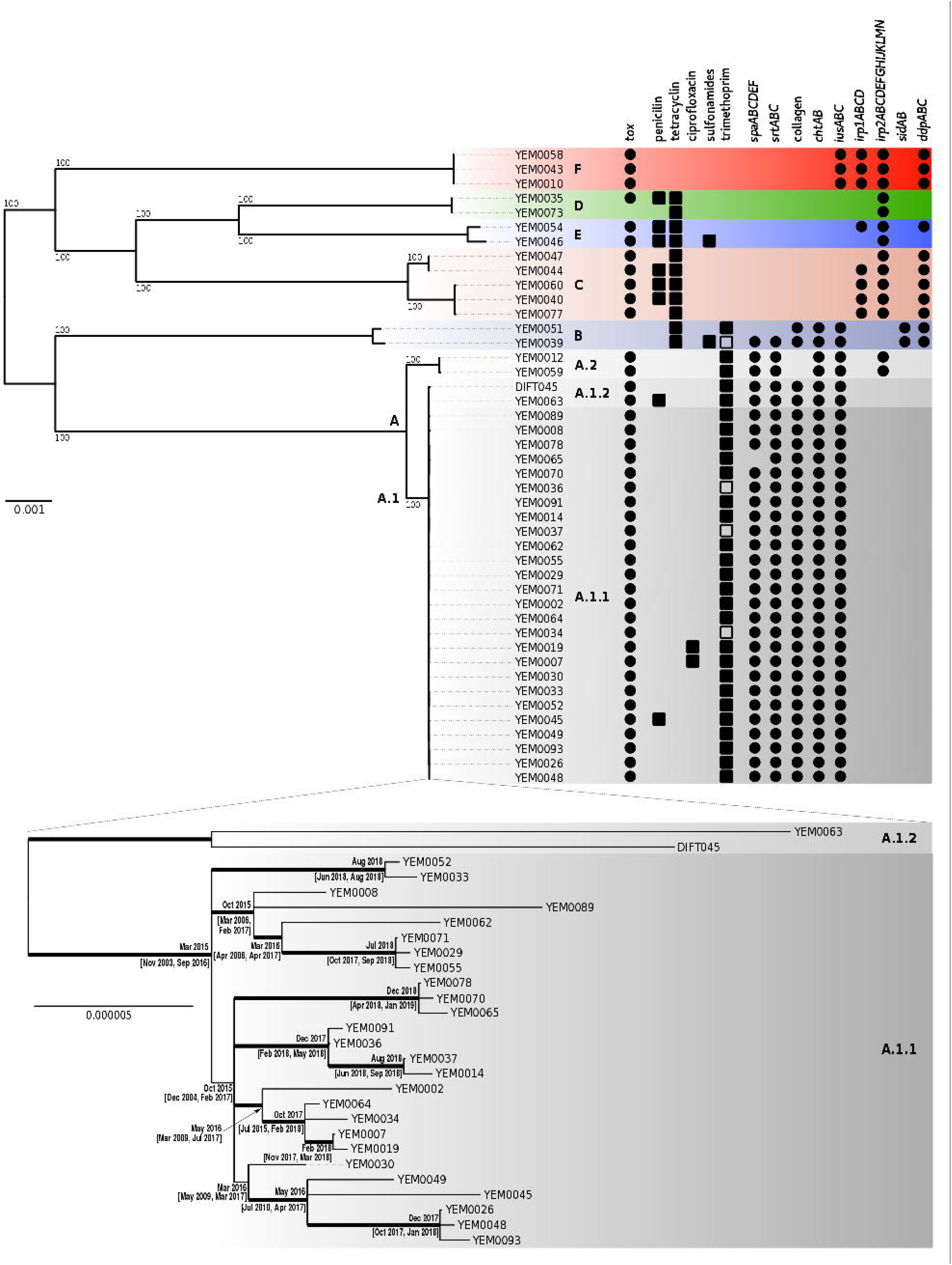
Phylogenetic analysis of Yemen isolates with their virulence and resistance characteristics. A. Phylogenetic tree of the Yemen isolates, with the distinction of 6 sublineages (A to F; sublineage A was subdivided into A.1 [A.1.1, A.1.2] and A.2). On the right of the tree, the black squares denote a resistance phenotype, while a black circle denote the presence of the *tox*-gene or other virulence-associated genes. Open squares denote an intermediate resistance phenotype. Scale bar, 0.001 nucleotide substitutions per site. B. Detailed phylogenetic tree of the sublineage A. Nodes ‘ages and 95%CI are shown at the nodes. Scale bar, 0.000005 nucleotide substitutions per site (*i.e*., 1 substitution every 200,000 nucleotides, or approximately 10 substitutions per genome).

Sublineages A and B were of biovar Gravis, whereas the others were biovar Mitis. The detection of the *spuA* gene, coding for a putative alpha-1,6-glycosidase [22] associated with the glycogen positive phenotype of biovar Gravis, was consistent with these biochemical assignations (**Figure 3A**) and showed that Yemen isolates belonged to two deep lineages of *C. diphtheriae* marked by the presence or absence of this gene (**Figure S2**) as recently reported [23].

Genotyping of the *C. diphtheriae* Yemen isolates by 7-gene multilocus sequence typing (MLST, **Table S1**) revealed that sublineage A isolates belonged to ST384. The single previously reported ST384 isolate (DIFT045; https://pubmlst.org/cdiphtheriae/) was isolated in Belgium in 2016, and was collected in a patient who had close contact with a person having traveled back from Saudi Arabia [24]. This observation hinted to a possible epidemiological link between the main Yemen outbreak lineage and Saudi Arabia. To reinforce this hypothesis, the genome of DIFT045 was sequenced, confirming its very close genetic relatedness with sublineage A (**Figure 3**). Based on the global phylogenetic tree (**Figure S2**), sublineage A was the Yemenite sublineage closest to the ex-Soviet Union 1990s outbreak strain [25]. However, the large genetic distance between sublineage A isolates and strain NCTC 13129, the reference for the ST8 ex-Soviet Union outbreak, rules out a direct epidemiological link between the two outbreaks.

Sublineage E isolates were closely related to two isolates collected from migrants from Somalia [26], and sublineage C isolates were closely related to isolates from siblings with a Somalian origin [7]. Other Yemen sublineages were not phylogenetically closely related to isolates from previously sequenced *C. diphtheriae* isolates (**Figure S2**).

We defined the genomic diversity of Yemen and DIFT045 isolates by clustering their protein-coding genes (**Figure S3**). A total of 3619 gene families were identified, 1790 of which were shared by all isolates (core genome). Genomes had 2277 genes on average (range 2234-2427), and each of the six sublineages possessed 54 to 140 genes not observed in the other sublineages, underlying the potential for large phenotypic variation among them.

### Microevolution of sublineage A

We subdivided sublineage A into two branches, with branch A.1 being further subdivided into A.1.1 and A.1.2 (**Figure 3**). The latter comprised the Belgium isolate DIFT045 and YEM0063. The epidemiologically predominant branch A.1 was highly homogeneous genetically, with a maximum of 86 genome-wide SNPs among its members (observed between YEM0063 and YEM0089), indicating that A.1 isolates share a recent common ancestor. The amount of nucleotide substitutions observed in isolates of this sublineage was significantly related to their isolation time (**Figure S4**), demonstrating measurable evolutionary divergence (Figure 3B), with a substitution rate estimated at 1.67 × 10^−6^substitutions per site per year (95%CI = 3.98 × 10 - 2.99 × 10). The last common ancestor of A.1.1 was estimated to have existed in March 2015 (Nov. 2003 - Sep. 2016). These results indicate that this subgroup of Yemenite diphtheria isolates circulated and diversified silently within the country several years before the diphtheria outbreak declaration in 2017.

### Antimicrobial susceptibility of Yemen *C. diphtheriae* isolates

Antimicrobial susceptibility data were heterogeneous among Yemen sublineages (**Figure 3, Table S2**). Importantly, full susceptibility or only low levels of penicillin resistance were observed for all Yemen diphtheria isolates. While eight isolates (18.6%) were penicillin resistant (**Table S2**), the two most resistant isolates had a minimal inhibitory concentration (MIC) of 0.25 mg/L, which is only twice the clinical breakpoint of resistance (>0.125 mg/L). Further, the MIC50 and MIC90 of penicillin were 0.094 and 0.19 mg/L, respectively (**Table S2**). Amoxicillin and erythromycin were also active against all isolates, as were many other tested molecules. Resistance to ciprofloxacin (but not to moxifloxacin) was observed in two isolates, which had a D93G change in the quinolone-resistance determining region of their *gyrA* gene. Resistance to trimethoprim was found in 32 (74.4%) isolates, including most members of sublineage A (**Figure 3**), but none was resistant to the combination of this agent with sulfamethoxazole (**Table S2**). Four isolates possessed homologs of the *sul1* gene, and decreased phenotypic susceptibility to sulfonamide was observed in two of these. Resistance to tetracycline was observed in 11 isolates, which all belonged to minor sublineages whereas sublineage A was susceptible (**Figure 3A**); five of these isolates possessed known tetracycline resistance genes encoding the *tet33* efflux pump or the *tetO* ribosomal protection protein. Finally, the two isolates of sublineage B possessed an *aadA2* gene coding for an ANT(3”) aminoglycoside modifying enzyme targeting spectinomycin and streptomycin. Accordingly, these two isolates were susceptible to gentamicin and kanamycin.

### Association of sublineage A with clinical signs and genomic features potentially linked to virulence

Clinical features of *C. diphtheriae* infections caused by isolates from sublineage A were distinctive, with patients tending to be younger (*p*=0.06) and with more frequent swollen neck (*p*=0.002; **Table 1**). We screened the genomic sequences for the presence of putative pathogenicity-associated genes. Sublineage A isolates possessed genes *iusABC* coding for an ABC-type iron uptake system, the *chtAB* genes that are homologous to *htaAB* genes for hemin binding, a pathogenicity island of *C. diphtheriae* called PICD-11 [27] comprising a putative collagen binding protein suggested to be associated with cutaneous or invasive infections [28], and the *spaABC-strA* and *spaDEF-strBC* gene clusters coding for two distinct types of fimbriae (**Figure 3**). In contrast to sublineage A, most other Yemen isolates did not harbor the above gene clusters. In turn, their genomes comprised genes *sidBA-ddpABC* for putative siderophore biosynthesis, *irp1ABCD* for a putative ABC-type iron transporter [29] and the *irp2ABCDEFGHI* and *irp2JKLMN* operons. Genes coding for SpaH fimbriae were absent from all Yemen genomes. Broader putative virulence genes variation was observed at the global scale, with a marked dichotomy between the two major lineages defined by gene *spuA* (**Figure S2**).

**Table 1.** Clinical characteristics of patients infected by sublineage A or other sublineages circulating in Yemen.

| Characteristics, (%) otherwise stated | Sublineage A<br>N=28 | Other sublineages<br>N=14 | P value* |
| --- | --- | --- | --- |
| Age, years, median (range) | 8 (2-35) | 14 (2-35) | 0.06 |
| Male | 14 (50) | 5 (36) | 0.38 |
| History of previous vaccination | 11 (44) | 7 (64) | 0.24 |
| Fever | 28 (100) | 14 (100) | - |
| Pseudomembrane | 28 (100) | 14 (100) |  |
| Neck swelling | 22 (84) | 4 (33) | 0.002 |

\* Differences between cases infected with *C. diphtheriae* sublineage A compared to the ones infected with isolates from other sublineages (B, C, D, E or F) were assessed by the Mann-Whitney U test, the Student's t test, the $\chi^2$ method, and Fisher's exact test where appropriate.

We further searched for sublineage A.1-enriched genes by analyzing the pangenome of Yemen isolates together with the sublineage A-related strain NCTC13129. 77 genes were enriched in sublineage A.1 (present in >95% A.1 isolates, and absent in >85% of other isolates), among which six were uniquely found in all members of this sublineage (**Table S4**). Among these, one gene was annotated as a 5-formyltetrahydrofolate cyclo-ligase gene, the homolog of which was previously shown to increase the resistance level to antifolates in *Mycobacterium smegmatis* and *Escherichia coli* [30]. Further analyses showed that this gene is in fact found in all *Corynebacterium* genomes, but the sublineage A.1 version is shorter (417 versus a median of 585 bp among all *Corynebacterium* species). Of note, the size of this gene does not vary at all in *C. diphtheriae* except for isolates of the Yemen outbreak sublineage A (585 bp). A possible causal link of this genetic truncation with resistance to trimethoprim, which was observed mostly in sublineage A (but not solely A.1; **Figure 3**), will be interesting to investigate in future studies.

## Discussion

The outbreak of diphtheria in Yemen was first notified in October 2017 and its early epidemiology was described [9, 11, 14]. Here, we provide an update on the epidemiological situation, a clinical description of the characteristics of recorded suspected cases, and detailed information on microbiological features of *C. diphtheriae* outbreak isolates.

Based on currently reported probable cases, the Yemen diphtheria outbreak is one of the largest *C. diphtheriae* contemporaneous outbreaks. We identified three discernible waves with distinctive geographic signatures. We note that reporting is very likely to be an underestimate in the context of the Yemen health care disruption, and to be uneven geographically given the heterogeneity in the continuation of health care facility operations. Nevertheless, the eDEWS system [31] proved invaluable in the surveillance of diphtheria in the conflict context.

Clinical characteristics were typical of diphtheria in terms of laryngeal manifestations, pseudomembrane presence and neck swelling [32]. Yemenite patients were mostly young, which is also typical, with the exception of the ex-Soviet Union event [4]. Vaccination is highly effective against the clinical expression of toxigenic *C. diphtheriae* infections, and countries with high population coverage have virtually eliminated endemic diphtheria [2]. In Yemen, the vaccination coverage of the third dose of pentavalent vaccine in 2017 and 2018 (**Figure S5**) was <80% in several governorates, which is considered as insufficient to reach population immunity [33]. We found that more than 50% of patients were reported to be previously vaccinated. We did not have access to the exact vaccination status, but we can assume that this observation reflects an incomplete vaccination without booster. This might also explain the median age > 5 years of patients, comparable to other outbreaks in populations where vaccination was incomplete [34][35]. From November 2017 to March 2018, a mass vaccination campaign targeted nearly 2.7 million children aged 6 weeks to 15 years in 11 governorates [36]. Here we found that from the first to the second wave, there was a reduction in infection risk in 0-4 years old, consistent with some success from the mass immunization program. In 2019, diphtheria vaccination was further conducted in 186 districts of the 12 Northern governorates [37], with 1.2 million children 6 weeks to 5 years of age vaccinated with the Penta vaccine and 2.2 million children 5-15 years with the Td vaccine. Vaccination against diphtheria in selected districts of Southern governorates and Sa ‘adah has started in July 2020. However, despite efforts to control the disease, cases were still being reported in 2020, and future studies will be needed to evaluate their effect.

This work represents a unique genomic analysis of a large contemporaneous outbreak of diphtheria. A prominent observation was that at least six phylogenetic sublineages, ancestrally unrelated within the global diversity of *C. diphtheriae*, contributed to the resurgence of diphtheria in Yemen. This important observation shows that strains with distinctive genomic and phenotypic characteristics can coexist within a single diphtheria outbreak. The multiplicity of strains reflects a silent diversity reservoir of *C. diphtheriae* despite vaccination, which is designed to protect against disease expression rather than colonization and transmission. Antimicrobial resistance remains rare in *C. diphtheriae* [38], and our results show that the main recommended agents (penicillin G, aminopenicillin and erythromycin) are active against Yemen *C. diphtheriae* despite a low-level penicillin resistance in some isolates. However, antimicrobial susceptibility profiles were heterogeneous, with a strong phylogenetic sublineage effect, and were largely concordant with genomic data, implying a possible future role for molecular testing of antimicrobial resistance in *C. diphtheriae*.

Our work also uncovers a large heterogeneity in virulence-associated genomic features among circulating isolates. The diphtheria toxin gene was observed in most isolates, which were all confirmed to produce the toxin *in vitro* except one, which was non-toxigenic toxin gene bearing (NTTB). However, three isolates were *tox*-negative, indicating that such isolates can cause diphtheria-like respiratory symptoms. Currently, with the important exception of diphtheria toxin, the links between virulence genes of *C. diphtheriae* strains and clinical expression are unknown [39]. Whereas two Yemen sublineages are of biovar Gravis, four were of biovar Mitis. Biovar Gravis isolates have been considered more virulent than biovar Mitis isolates [40, 41]. Interestingly, our work reveals that the distribution of virulence-associated genes is strongly contrasted between two major lineages of *C. diphtheriae*, which were defined here by the presence of the Gravis-specific gene *spuA*. Our genomic data suggest that inter-lineage heterogeneity in iron acquisition, adhesion and colonization capacities may exist and could underlie epidemiological or clinical differences among them. We found that patients infected with sublineage A, of biovar Gravis, tended to be younger and with more frequent neck swelling. However, as neck swelling is more commonly observed in young patients, we cannot disentangle sublineage and age effects here. The underlying mechanisms of lymph node manifestations in diphtheria, described as nonspecific acute lymphadenitis [1], are unknown. Future studies should investigate the impact of the genomic diversity of *C. diphtheriae* on the pathogenicity and clinical expression of diphtheria.

Genomic sequencing of bacterial pathogens is a powerful approach to define relationships across time, sources and geography [42][15]. The *C. diphtheriae* genotypes observed in Yemen were distinct from strains from the large ex-Soviet Union 1990s outbreak, whereas no data exists on the strains involved in other recent outbreaks in Venezuela, Haiti and Bangladesh. Migration and trade from the horn of Africa into Yemen are major drivers of pathogenic strain spread, as observed for cholera [15]. The nearly complete lack of information on circulating *C. diphtheriae* from neighboring countries has restricted our ability to define the dynamics of geographic spread of *C. diphtheriae* in this world region. However, a few *C. diphtheriae* genomes were linked epidemiologically and genetically to the neighboring countries Somalia and Saudi Arabia.

We estimated the evolutionary rate of *C. diphtheriae* and found it is similar to other human pathogens sampled over short time scales, such as *Staphylococcus aureus* or *Streptococcus pyogenes* [43]. Our genomic data analyses thus imply that the diversification of the major Yemen outbreak sublineage largely predates the detection of the outbreak by the disease surveillance system. These observations concur with the multiplicity of sublineages that a reservoir of *C. diphtheriae* diversity existed in Yemen before the outbreak recognition. We note that our rate and age estimates have large confidence intervals, which is explained by the small number of available genomes.

Although we provide a glimpse into the microbiological characteristics of diphtheria in Yemen, the available samples of *C. diphtheriae* sent for biological confirmation currently do not provide a complete picture of the outbreak. Samples referred to NCPHL were distributed across a large time period but governorates of the Southern part of the country were largely under-represented, as 91% of confirmed samples came from Northern governorates (**Table S3**). This limitation reflects the current situation of the country, divided by conflict into South and North parts.

In summary, this work sheds light on epidemiological and clinical aspects of the current Yemen diphtheria outbreak. We demonstrate high phylogenetic, genomic and phenotypic variation of *C. diphtheriae* within a single outbreak, suggesting that the characterization of isolates at the level of individual patients or local chains of transmission is relevant for patient management during diphtheria outbreaks. This underlines the need to reinforce laboratory capacity in the context of diphtheria outbreaks. Concerted international efforts should be developed aiming at enhancing surveillance and defining in real time the characteristics of *C. diphtheriae* clinical isolates. Diphtheria is a largely forgotten disease, and contemporaneous research into its pathophysiology is scarce. Further, in face of increasing disruption of vaccination campaigns and the shortage of diphtheria antitoxin supply, studies into the determinants of local persistence of *C. diphtheriae* and its spread at regional or global scales are needed in order to better control the reemergence of this dreadful pathogen.

## Methods

### Definitions and surveillance data

Probable cases were defined based on clinical examination: any person with illness characterized by an adherent membrane on the tonsils, pharynx and/or nose and any one of the following: laryngitis, pharyngitis or tonsillitis [31]. Confirmed cases were defined as a probable case from which a *C. diphtheriae* isolate was cultivated in the NCPHL laboratory. Probable cases were based on the health facility surveillance system and were compiled from electronic diseases early warning system (eDEWS) releases of weekly reporting of diphtheria cases [31]. For the vaccination coverage, we used the 2017 and 2018 annual reports by the national Expanded Programme on Immunization (EPI), obtained from the Ministry of Public Health and Population (MoPHP) of Yemen. The attack rate per age group was calculated by dividing the number of cases of each age group by the size of the population within each age group (source: https://www.census.gov/data-tools/demo/idb/informationGateway.php).

### Biological confirmation, clinical data and external microbiological and genomic analyses

A total of 888 throat swab samples were received by the NCPHL and analyzed using local microbiological procedures (Supplementary data). Demographic data, prior vaccination, signs and symptoms and current antibiotic of patients were collected for all samples. A random selection of 98 samples available on July 2019 was sent to Institut Pasteur for external analyses (Supplementary data).

## Data Availability

The whole genome sequencing data generated in this study were deposited in the European Nucleotide Archive (ENA) database and are accessible through the study PRJEB34206 (https://www.ebi.ac.uk/ena/data/view/PRJEB34206). Raw read data are available under the accessions ERR4332853-96. Contig sequences are available under the accessions CAJDXH01000000-CAJDYY01000000.

## Ethical statement

Data were collected as part of routine case management under an emergency response mandate from the government of Yemen. The scientific committee of the NCPHL, Sana ‘a authorized the usage of the clinical data included in this study.

## Author contributions

A.H., the NCPHL diphtheria outbreak working group (A.A.R, M.A.A., E.M.A.-A., N.M.A.-M., K.A.A, H.Z. A.-S., A.A.A.-S.), A.C-L., E.B., M.P.P. and M.L. performed the microbiological cultures of the isolates and their biochemical and molecular characterizations. F.D., G.D., N.L. and H.S. collated and analyzed the epidemiological data. A.H., G.D. and J.T. collated and analyzed the clinical data. A.C., V.B., M.H., J.G., N.Z. and S.B. analyzed the genomic data. H.M. provided the DIFT045 strain. S.B. and G. D. initially designed, and subsequently coordinated, the project. S.B. designed the microbiological aspects of the study and wrote the initial version of the manuscript. All authors provided input to the manuscript and reviewed the final version.

## Acknowledgments

We thank Dr Taha Almotuakel (Yemen Ministry of Health) for continuous support, and Isabelle Cailleau (Institut Pasteur) and the Medecins Sans Frontieres Organization for logistical support. The NCPHL also acknowledges the World Health Organization for providing some reagents used in this study. This work used the computational and storage services (TARS cluster) provided by the IT department at Institut Pasteur.

## References

1. Hadfield TL, McEvoy P, Polotsky Y, Tzinserling VA, Yakovlev AA. The pathology of diphtheria. J Infect Dis 2000;181 Suppl 1:S116–120.

2. Sharma NC, Efstratiou A, Mokrousov I, Mutreja A, Das B, et al. Diphtheria. Nat Rev Dis Primer 2019;5:81.

3. Patey O, Bimet F, Riegel P, Halioua B, Emond JP, et al. Clinical and molecular study of Corynebacterium diphtheriae systemic infections in France. Coryne Study Group. J Clin Microbiol 1997;35:441–445.

4. Dittmann S, Wharton M, Vitek C, Ciotti M, Galazka A, et al. Successful control of epidemic diphtheria in the states of the Former Union of Soviet Socialist Republics: lessons learned. J Infect Dis 2000;181 Suppl 1:S10–22.

5. Rahman MR, Islam K. Massive diphtheria outbreak among Rohingya refugees: lessons learnt. J Travel Med;6. Epub ahead of print 1 January 2019. DOI: 10.1093/jtm/tay122.

6. Paniz-Mondolfi AE, Tami A, Grillet ME, Márquez M, Hernández-Villena J, et al. Resurgence of Vaccine-Preventable Diseases in Venezuela as a Regional Public Health Threat in the Americas. Emerg Infect Dis 2019;25:625–632.

7. Meinel DM, Kuehl R, Zbinden R, Boskova V, Garzoni C, et al. Outbreak investigation for toxigenic Corynebacterium diphtheriae wound infections in refugees from Northeast Africa and Syria in Switzerland and Germany by whole genome sequencing. Clin Microbiol Infect Off Publ Eur Soc Clin Microbiol Infect Dis 2016;22:1003.e1-1003.e8.

8. Scheifer C, Rolland-Debord C, Badell E, Reibel F, Aubry A, et al. Re-emergence of Corynebacterium diphtheriae. Med Mal Infect 2019;49:463–466.

9. Dureab F, Müller O, Jahn A. Resurgence of diphtheria in Yemen due to population movement. J Travel Med;25. Epub ahead of print 1 January 2018. DOI: 10.1093/jtm/tay094.

10. Dureab F, Al-Sakkaf M, Ismail O, Kuunibe N, Krisam J, et al. Diphtheria outbreak in Yemen: the impact of conflict on a fragile health system. Confl Health 2019;13:19.

11. WHO. Diphtheria – Yemen. Disease Outbreak News. https://www.who.int/csr/don/22-december-2017-diphtheria-yemen/en/ (2017).

12. OCHA. Yemen: Diphtheria Outbreak -Nov 2017. https://reliefweb.int/disaster/ep-2017-000175-yem (2017).

13. Meyer D. Yemen--First cholera, now diphtheria. https://www.outbreakobservatory.org/outbreakthursday-1/11/30/2017/yemen-first-cholera-now-diphtheria (2017).

14. WHO-EMRO. Weekly Epidemiological Monitor, Volume 10, Issue 47 (19 November 2017). https://reliefweb.int/report/yemen/who-emro-weekly-epidemiological-monitor-volume-10-issue-47-19-november-2017 (2017).

15. Weill F-X, Domman D, Njamkepo E, Almesbahi AA, Naji M, et al. Genomic insights into the 2016-2017 cholera epidemic in Yemen. Nature 2019;565:230–233.

16. Dureab F, Al-Awlaqi S, Jahn A. COVID-19 in Yemen: preparedness measures in a fragile state. Lancet Public Health 2020;5:e311.

17. Dhabaan GN, Al-Soneidar WA, Al-Hebshi NN. Challenges to testing COVID-19 in conflict zones: Yemen as an example. J Glob Health 2020;10:010375.

18. eDEWS. Electronic Integrated Disease Early Warning and Response System, Volume 08, lssue19, Epi week 19,(04-10 May,2020).

19. Wagner KS, Stickings P, White JM, Neal S, Crowcroft NS, et al. A review of the international issues surrounding the availability and demand for diphtheria antitoxin for therapeutic use. Vaccine 2009;28:14–20.

20. Kneen R, Pham NG, Solomon T, Tran TM, Nguyen TT, et al. Penicillin vs. erythromycin in the treatment of diphtheria. Clin Infect Dis Off Publ Infect Dis Soc Am 1998;27:845–850.

21. Badell E, Guillot S, Tulliez M, Pascal M, Panunzi LG, et al. Improved quadruplex real-time PCR assay for the diagnosis of diphtheria. J Med Microbiol 2019;68:1455–1465.

22. Santos AS, Ramos RT, Silva A, Hirata R, Mattos-Guaraldi AL, et al. Searching whole genome sequences for biochemical identification features of emerging and reemerging pathogenic Corynebacterium species. Funct Integr Genomics 2018;18:593–610.

23. Hennart M, Panunzi LG, Rodrigues C, Gaday Q, Baines SL, et al. Population genomics and antimicrobial resistance in Corynebacterium diphtheriae. bioRxiv 2020;2020.05.19.101030.

24. Martini H, Soetens O, Litt D, Fry NK, Detemmerman L, et al. Diphtheria in Belgium: 2010-2017. J Med Microbiol 2019;68:1517–1525.

25. Cerdeño-Tárraga AM, Efstratiou A, Dover LG, Holden MTG, Pallen M, et al. The complete genome sequence and analysis of Corynebacterium diphtheriae NCTC13129. Nucleic Acids Res 2003;31:6516–6523.

26. Berger A, Dangel A, Schober T, Schmidbauer B, Konrad R, et al. Whole genome sequencing suggests transmission of Corynebacterium diphtheriae-caused cutaneous diphtheria in two siblings, Germany, 2018. Euro Surveill Bull Eur Sur Mal Transm Eur Commun Dis Bull;24. Epub ahead of print January 2019. DOI: 10.2807/1560-7917.ES.2019.24.2.1800683.

27. Trost E, Tauch A. Comparative genomics and pathogenicity islands of Corynebacterium diphtheriae, Corynebacterium ulcerans, and Corynebacterium pseudotuberculosis. In: Corynebacterium diphtheriae and related toxigenic species: genomics, pathogenicity and applications. Burkovski, A. Ed.; Springer; 2014. pp. 39–65.

28. Timms VJ, Nguyen T, Crighton T, Yuen M, Sintchenko V. Genome-wide comparison of Corynebacterium diphtheriae isolates from Australia identifies differences in the Pan-genomes between respiratory and cutaneous strains. BMC Genomics 2018;19:869.

29. Qian Y, Lee JH, Holmes RK. Identification of a DtxR-regulated operon that is essential for siderophore-dependent iron uptake in Corynebacterium diphtheriae. J Bacteriol 2002;184:4846– 4856.

30. Ogwang S, Nguyen HT, Sherman M, Bajaksouzian S, Jacobs MR, et al. Bacterial conversion of folinic acid is required for antifolate resistance. J Biol Chem 2011;286:15377–15390.

31. Dureab F. The usefulness of the electronic Disease Early Warning System (eDEWS) in the humanitarian crisis of Yemen. University of Heidelberg. https://archiv.ub.uni-heidelberg.de/volltextserver/27986/ (2019).

32. Dash N, Verma S, Jayashree M, Kumar R, Vaidya PC, et al. Clinico-epidemiological profile and predictors of outcome in children with diphtheria: a study from northern India. Trop Doct 2019;49:96–101.

33. Blumberg LH, Prieto MA, Diaz JV, Blanco MJ, Valle B, et al. The Preventable Tragedy of Diphtheria in the 21st Century. International journal of infectious diseasesfZ: IJIDfZ: official publication of the International Society for Infectious Diseases;71. Epub ahead of print June 2018. DOI: 10.1016/j.ijid.2018.05.002.

34. Murhekar M. Epidemiology of Diphtheria in India, 1996-2016: Implications for Prevention and Control. Am J Trop Med Hyg 2017;97:313–318.

35. Galazka A. The changing epidemiology of diphtheria in the vaccine era. J Infect Dis 2000;181 Suppl 1:S2–9.

36. OCHA-services. Diphtheria vaccination campaign for 2.7 million children concludes in Yemen. https://reliefweb.int/report/yemen/diphtheria-vaccination-campaign-27-million-children-concludes-yemen (2018).

37. WHO-EMRO. Situation Report, September 2019, Issue NO.9: Yemen Conflict. https://reliefweb.int/sites/reliefweb.int/files/resources/Yem-Sitrep-Sept-2019.pdf (2019).

38. Zasada AA. Antimicrobial susceptibility and treatment. In: Corynebacterium diphtheriae and related toxigenic species genomics, pathogenicity and applications. Burkovski A., editor. New York: Springer; 2014. pp. 239–246.

39. Sangal V, Hoskisson PA. Evolution, epidemiology and diversity of Corynebacterium diphtheriae: New perspectives on an old foe. Infect Genet Evol J Mol Epidemiol Evol Genet Infect Dis 2016;43:364–370.

40. McLeod JW. THE TYPES MITIS, INTERMEDIUS AND GRAVIS OF CORYNEBACTERIUM DIPHTHERIAE:A Review of Observations during the Past Ten Years. Bacteriol Rev 1943;7:1–41.

41. Barksdale L. Corynebacterium diphtheriae and its relatives. Bacteriol Rev 1970;34:378–422.

42. Moura A, Criscuolo A, Pouseele H, Maury MM, Leclercq A, et al. Whole genome-based population biology and epidemiological surveillance of Listeria monocytogenes. Nat Microbiol 2016;2:16185.

43. Duchêne S, Holt KE, Weill F-X, Le Hello S, Hawkey J, et al. Genome-scale rates of evolutionary change in bacteria. Microb Genomics 2016;2:e000094.

